# Symptoms of depression can be more frequent in non-surgical patients with left lateralization of Temporal Lobe Epilepsy: A systematic review

**DOI:** 10.1101/2020.08.20.20178293

**Authors:** Graciane Radaelli, Fernanda Majolo, Eduardo Leal-Conceição, Francisco S. Santos, Vinicius S. Escobar, Gislaine Baroni, Gabriele G. Zanirati, Mirna W. Portuguez, Fulvio A. Scorza, Jaderson C. da Costa

## Abstract

Considering that the side of epileptogenic focus is a factor that could contribute to depressive and anxiety symptoms, we propose a systematic review searching for the prevalence of depression in Temporal Lobe Epilepsy in non-surgical patients. We performed a literature search in PubMed/Medline, Web of Science and PsycNET for data from inception until January 2019. The terms “epilepsy, temporal lobe OR “epilepsy” AND “temporal” AND “lobe” OR “temporal lobe epilepsy” OR “temporal” AND “lobe” AND “epilepsy” AND “depressive disorder” OR “depressive” AND “disorder” OR “depressive disorder” OR “depression” OR “depression” OR “anxiety” OR “anxiety” were used in the search strategy. After screening titles and abstracts, only 32 articles met the inclusion criteria. DSM/SCID is the main method utilized to psychiatric diagnosis. The majority of the studies did not perform neuropsychological evaluation. From 24 studies, most clinic cases of lateralization of epileptic focus depression symptoms showed lateralization in the left hemisphere. Nine studies were evaluated for individual depressive diagnosis, therefore, the analyzed data does not present statistical significance between right and left hemispheres. This study shows mood disorders are prevalent in epileptic patients undergoing clinical treatment. However, to date there is no correlation between lateralization of epilepsy and the prevalence of mood disorders or cognitive impairment. Well-conducted studies are needed to establish the correlation between the epilepsy lateralization and mood disorders.

## Introduction

Temporal Lobe Epilepsy (TLE) is the most common form of epilepsy and it is often refractory to antiepileptic drugs (AEDs), with variation of 53–76% of patients that are resistant to medical treatment [1–3]. TLE is highly associated with psychiatric comorbidities, and primarily mood, depression and anxiety disorders [4–8]. There is no consensus as to why this association exists, but the major agreement comes from the possibility that these comorbidities and TLE share similar neuroanatomic localizations [9–11]. Also, clinical and experimental studies evidenced neurobiological mechanisms that could linkage epilepsy and depression symptoms, such as differences in neurotransmitters [12] and alterations in brain glucose metabolism and metabolic network in regions related to the pathophysiology of both epilepsy and depression [13]. Another factor that could contribute to depressive and anxiety symptoms in TLE is the hemisphere of epileptogenic focus, but this still remains uncertain, with studies pointing to prevalence of interictal depression in left-sided seizure foci [14, 15], while others showing a tendency for greater depressive symptoms in presurgical right-sided seizure foci patients [14, 15].The most frequent way to evaluate depressive and anxiety symptoms in epilepsy patients is by using scales and inventories that provide a quantitative measure of recurrent and severity of mood symptomatology experienced by the patients [16]. The effects of underlying injury, mood disorders and AEDs treatment can also impact neuropsychological functions in TLE patients. In this way, neuropsychological assessment is substantial to better understand the impact of this variables in patients’ lives. There is no clear data of the interaction between laterality of temporal lobe epilepsy, usage of AEDs, depressive symptoms and neuropsychological functions. To better understand this relation, the aim of this study is to review the outcomes of the association between laterality of epileptogenic focus and its relationship with depressive symptoms in non-surgical patients.

## Methods

A systematic review using the methodology outlined in the Cochrane Handbook for Systematic Reviewers was performed [17]. The data were reported following the Preferred Reporting Items for Systematic Reviews and Meta-Analyses [18]. The review protocol was registered in the International Register of Prospective Systematic Reviews under the registration number CRD42019104443.

### Database search

A literature search was performed in the PubMed/Medline, Web of Science and PsycNET for data from inception until January 2019. The following terms and medical subject headings (MeSH) were used in the search strategy: (“epilepsy, temporal lobe OR “epilepsy” AND “temporal” AND “lobe” OR “temporal lobe epilepsy” OR “temporal” AND “lobe” AND “epilepsy” AND “depressive disorder” OR “depressive” AND “disorder” OR “depressive disorder” OR “depression” OR “depression” OR “anxiety” OR “anxiety”. The strategies for other databases are available on request. Articles published in all languages were included. The bibliography of the included articles was manually searched. Two authors (E.L.C. and F.S.S.) independently evaluated the titles and abstracts of all studies identified in the search based on the abovementioned terms and MeSH. Disagreements were resolved by consensus or by a third reviewer (G.R.).

### Eligibility criteria

The inclusion criteria were the following: articles without language restriction; diagnosis of epilepsy by ECG or MRI; Psychiatric diagnosis of depression (by scales, interviews, diagnostic manuals - DSM and ICD –10); study design (case series with more than 10 patients, retrospective and prospective, clinical, human); unilateral or bilateral temporal lobe epilepsy; non-surgical patients.

Exclusion criteria were studies of systematic reviews, letters, and experimental studies; children under 16; patients with dysphoria; patients with generalized epilepsy (multiple foci); surgical patients. Figure 1 shows a flowchart of study selection and inclusion.

**Figure 1.**
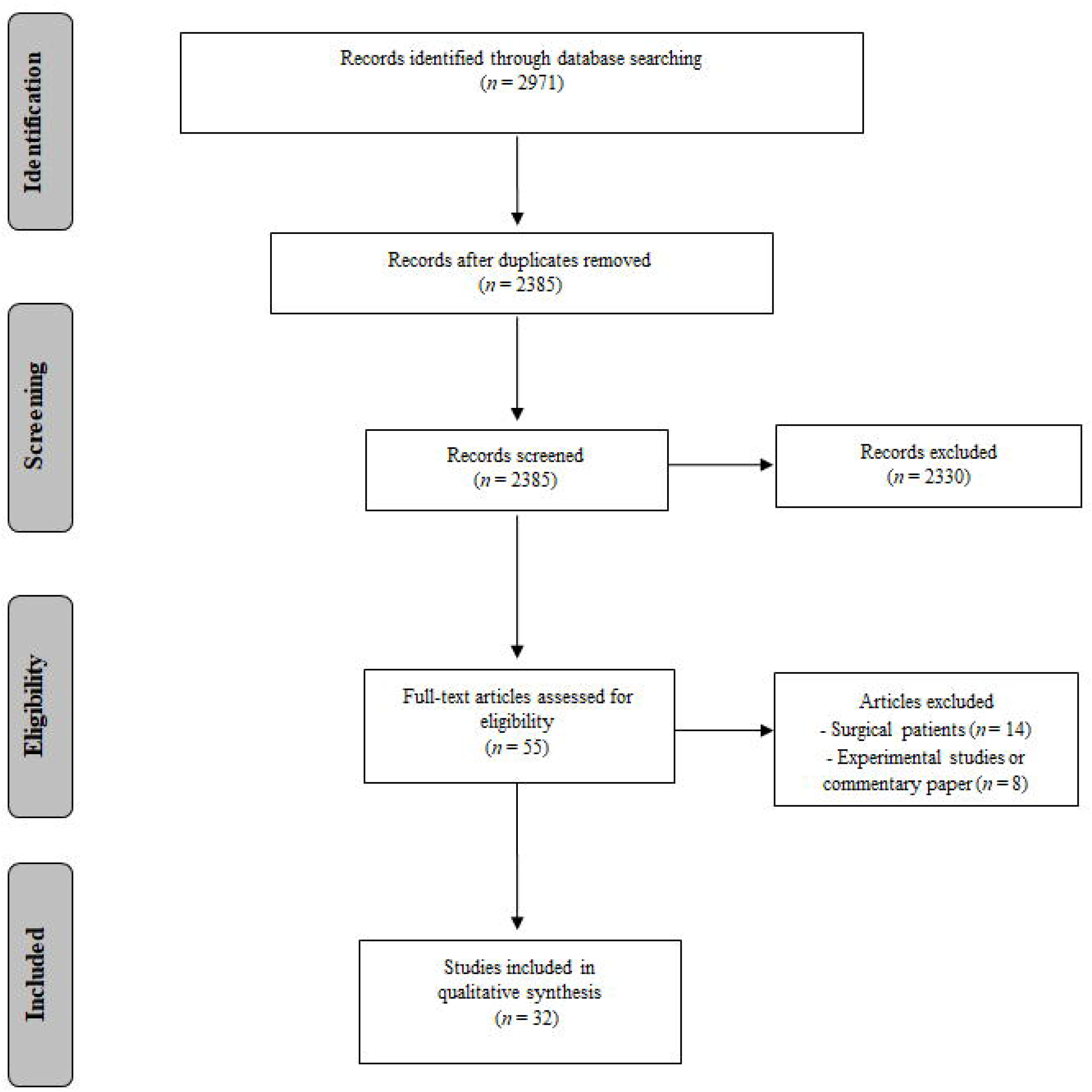
Summary of evidence search and study selection.

### Data extraction

The databases were searched and duplicate entries were removed. Abstracts that did not provide sufficient information regarding the inclusion and exclusion criteria were selected for full-text evaluation. After, the same reviewers independently evaluated the full text of these articles and made their selection in accordance with the eligibility criteria. Data on the following were collected: The number of patients, method of psychiatric diagnosis, symptoms diagnosis criteria, diagnostic scales, neuropsychological tests, report of the number of antiepileptic drugs, lateralization of epileptic focus, individual depression symptoms, lateralized depression symptoms, individual anxiety symptoms, lateralized anxiety symptoms and the design/classification of evidence.

## Results

The search retrieved 2971 potentially relevant citations from the electronic databases. Duplicate titles were removed, leaving 55 articles. After screening titles and abstracts, only 32 articles met the inclusion criteria (Fig. 1). Of all the performed studies, only four were multicenter [15, 19–21], all the others were single center and cross-sectional.

At the total, 32 non-surgical studies with TLE were evaluated, being DSM/SCID the main method utilized to psychiatric diagnosis (46.9%) and 13 studies did not perform psychiatric diagnosis(40.6%). Also, the majority of the studies (65.6%) did not perform neuropsychological evaluation. From 24 studies, most clinic cases of lateralization of epileptic focus depression symptoms showed lateralization in the left hemisphere (1201 patients) (Table 1). Nine studies were evaluated for individual depressive diagnosis, therefore, the analyzed data does not present statistical significance between right and left hemispheres (Table 2).

**Table 1.** Clinical characteristics of non-surgical patients with Temporal Lobe Epilepsy related to the diagnostic of depression and treatment.

| Study | N/(%) |
| --- | --- |
| **Sample (n = 32)** |  |
| Mean (SD) | 112.53 (±108.30) |
| **Design** |  |
| Cross-sectional | 32 (100) |
| **Trail** |  |
| Single center | 4 (12.5) |
| Multi center | 28 (87.5) |
| **Method of Psychiatric Diagnosis** |  |
| DSM/SCID | 15 (46.87) |
| ICD – 10 | 2 (6.25) |
| Other interview | 2 (6.25) |
| Symptom of depression / Scale | 13 (40.62) |
| **Cognitive Evaluation** |  |
| Yes | 11 (34.4) |
| No | 21 (65.6) |
| **Lateralization of epileptic focus (n = 24)** |  |
| Right | 1016 |
| Left | 1201 |
| Bilateral | 129 |
| Extra-temporal | 97 |
| **Lateralization of Depression Symptoms (n =9)** |  |
| Right | 92 |
| Left | 106 |
| Bilateral | 03 |
| **Antiepileptic Drugs** |  |
| Monotherapy | 187 |
| Polytherapy | 415 |
| Unknown | 70 |

*DSM, Diagnostic and Statistic Manual; ICD-10, International Classification of Diseases-10.*

**Table 2.**
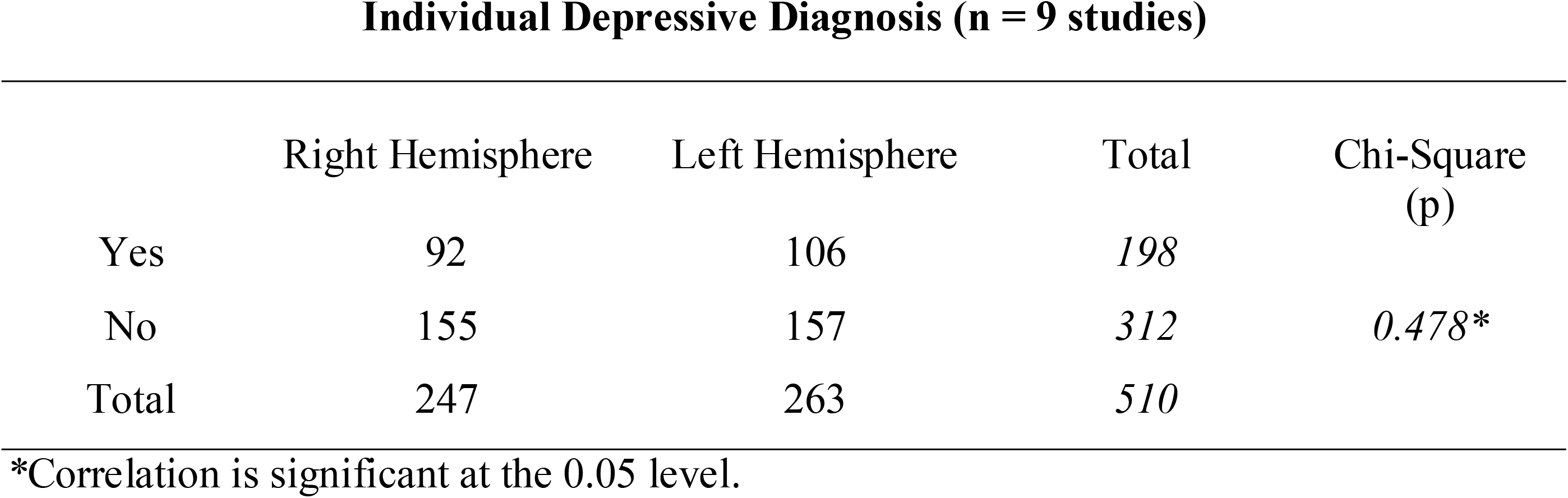
Clinical characteristics related to the lateralization of epileptic focus and depression symptoms of non-surgical patients with Temporal Lobe Epilepsy.

In regard to the kind of treatment using antiepileptic drugs, most of the patients utilized polytherapy (415 patients) instead of monotherapy (187 patients). More details about each evaluated study above can be visualized in Tables 3 and 4.

**Table 3.**
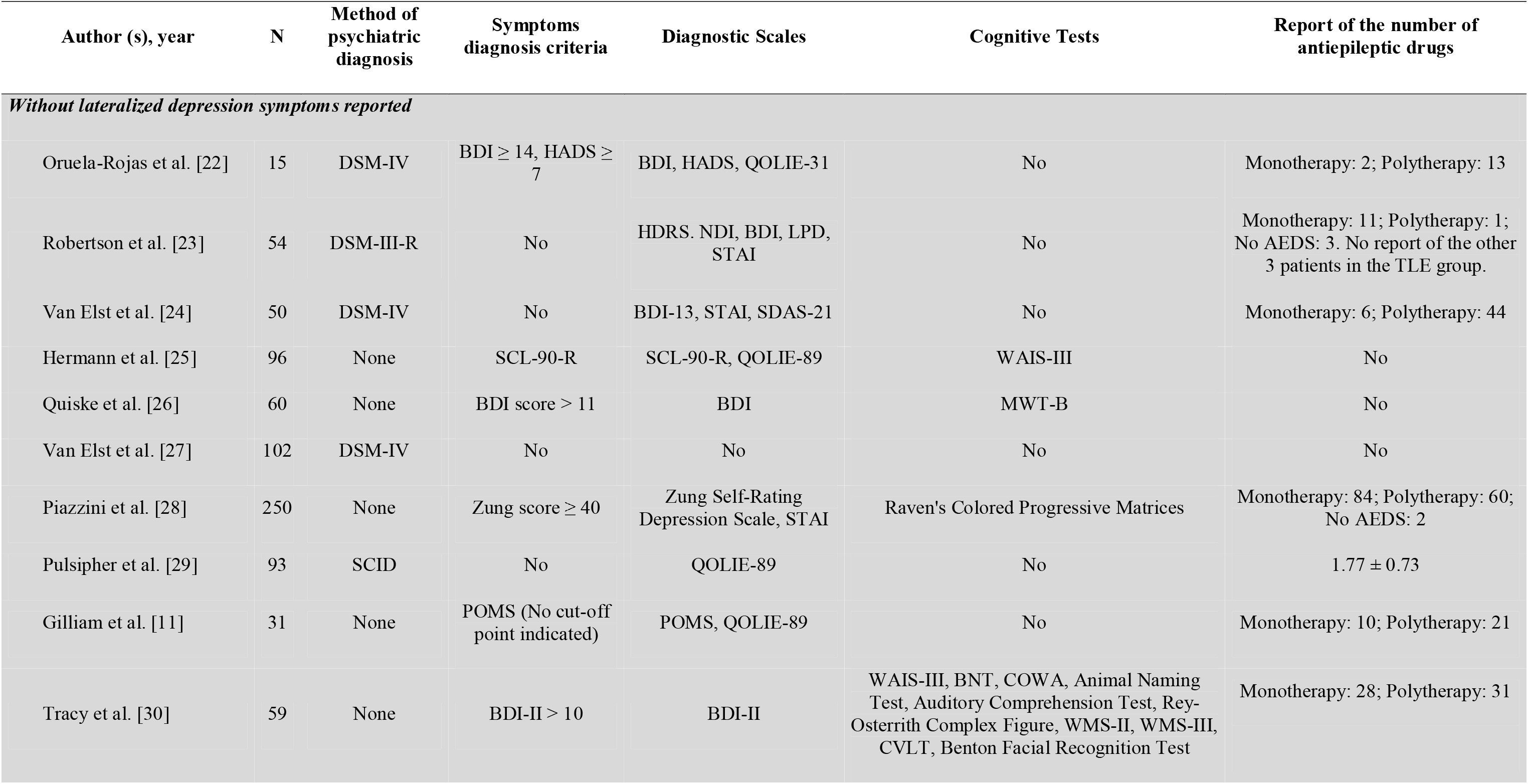
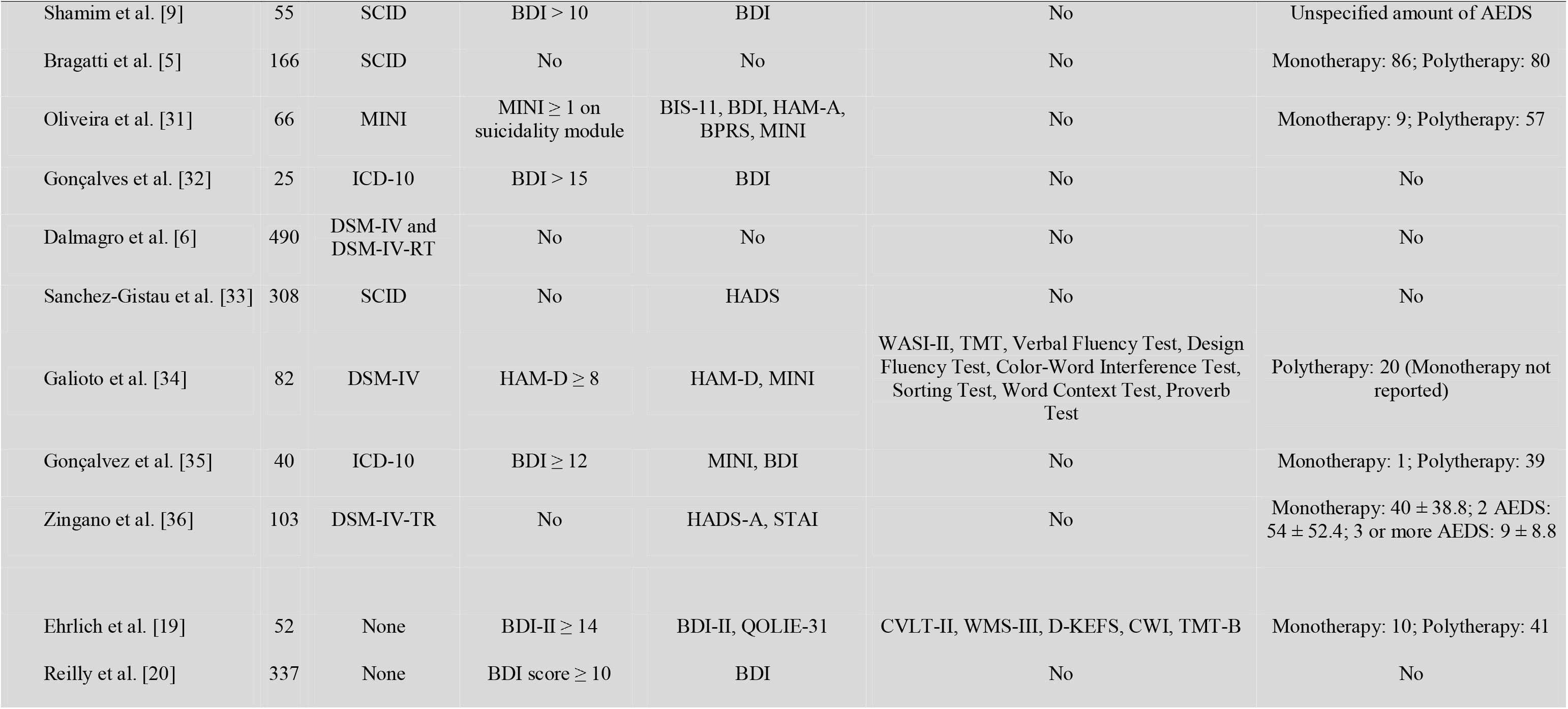
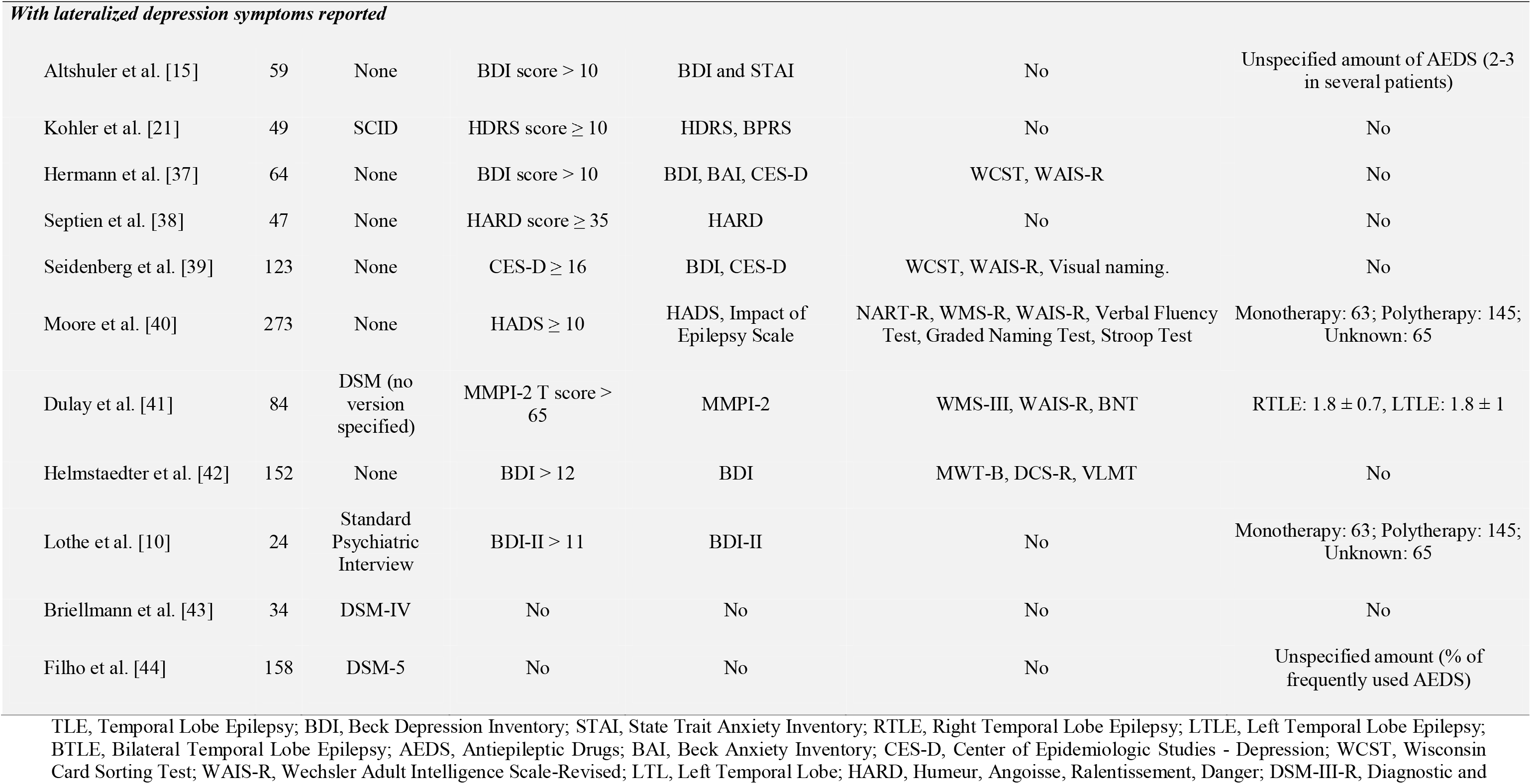
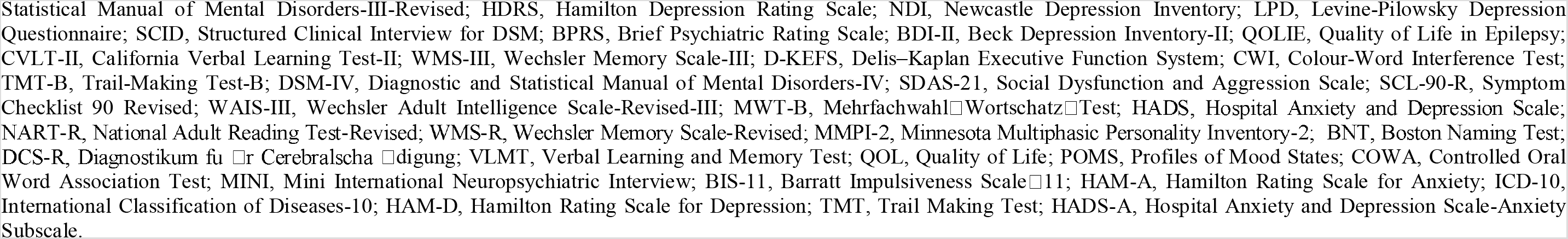
Clinical characteristics related to the diagnostic of depression and treatment of non-surgical patients with Temporal Lobe Epilepsy.

**Table 4.**
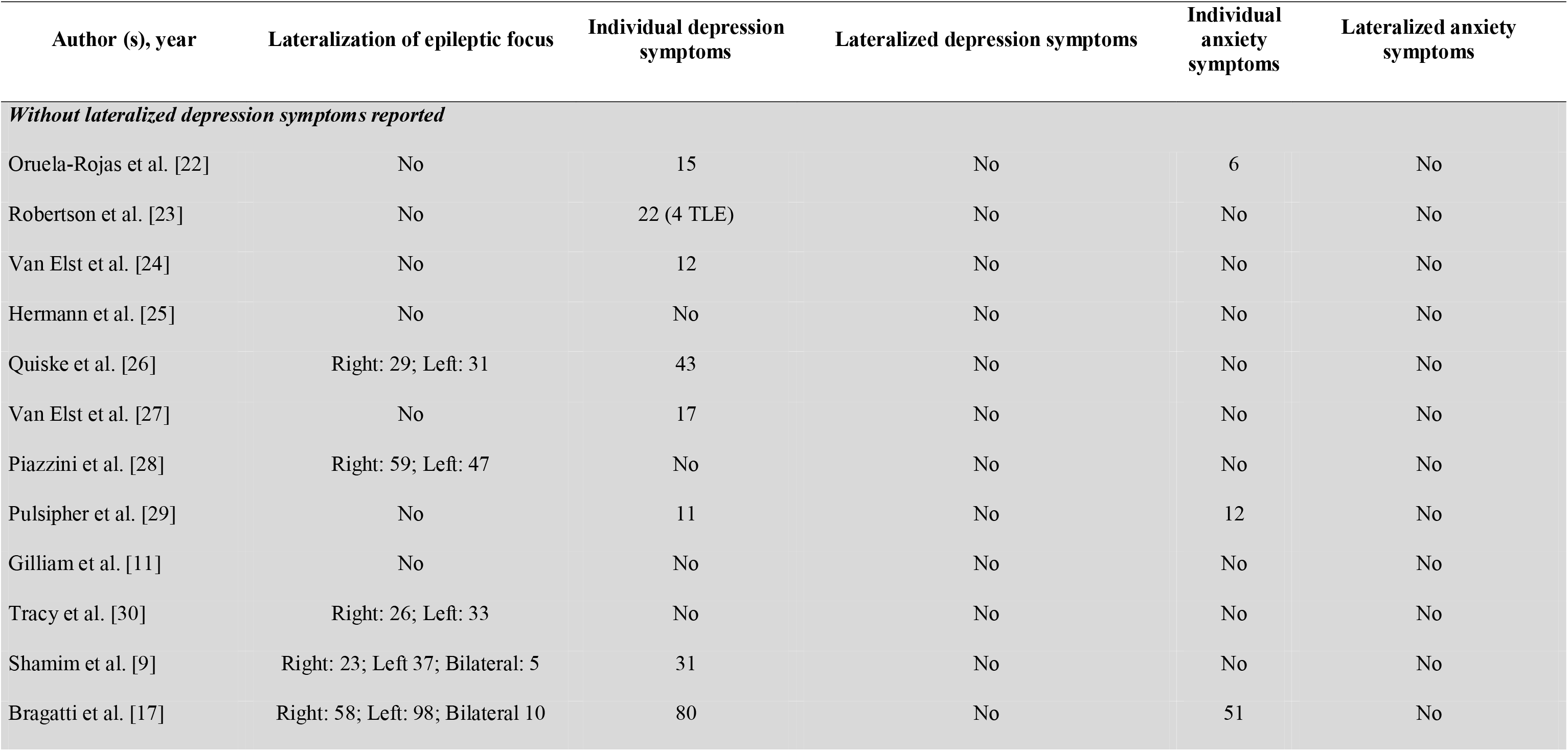
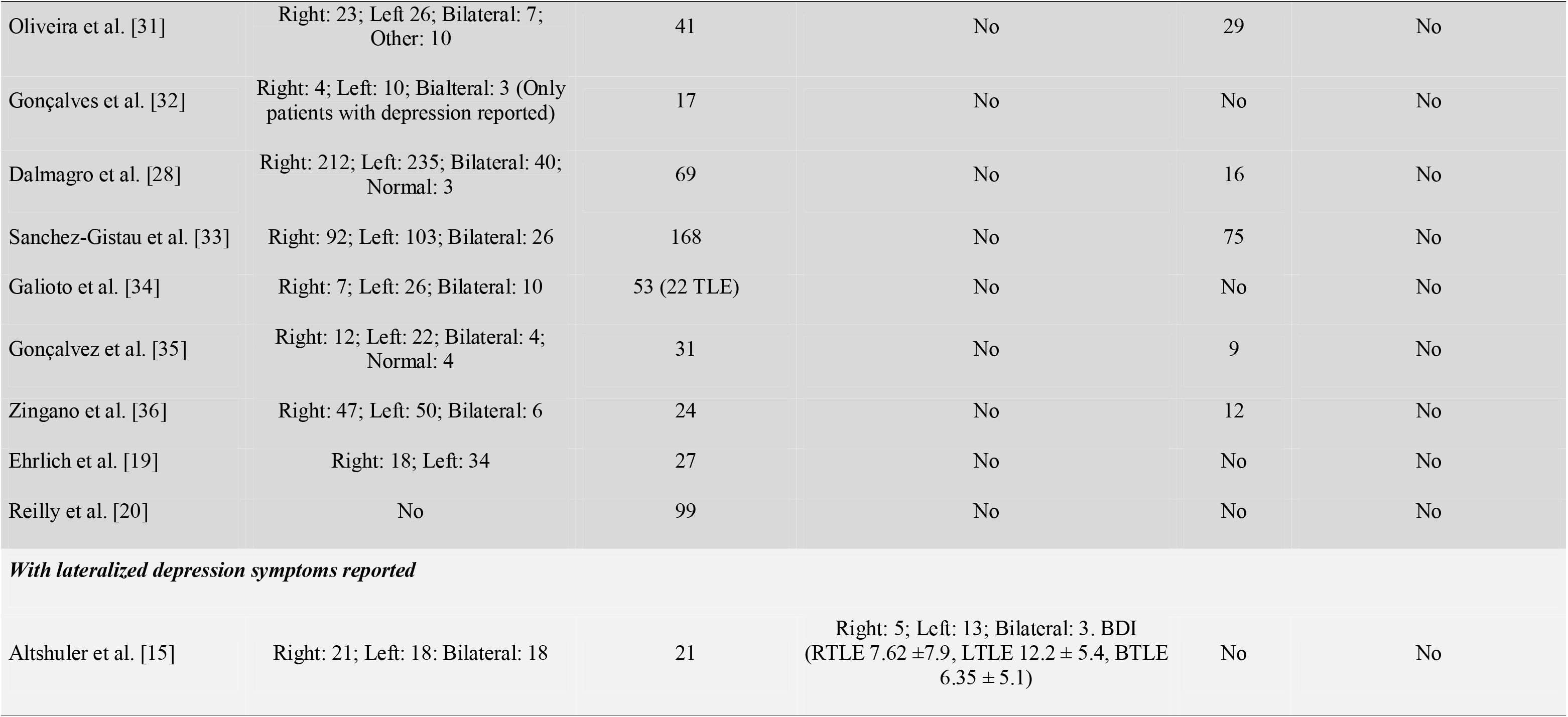
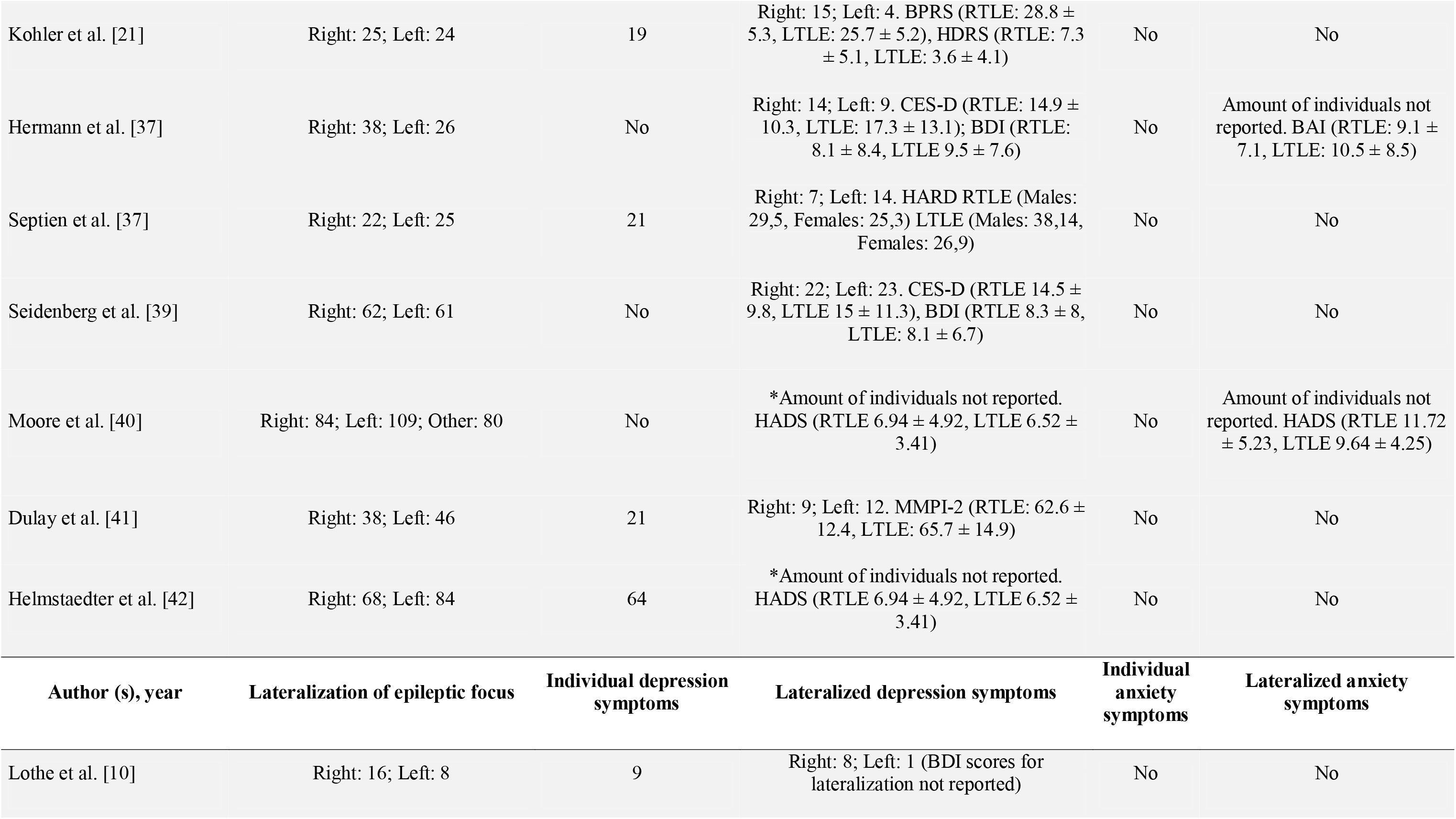
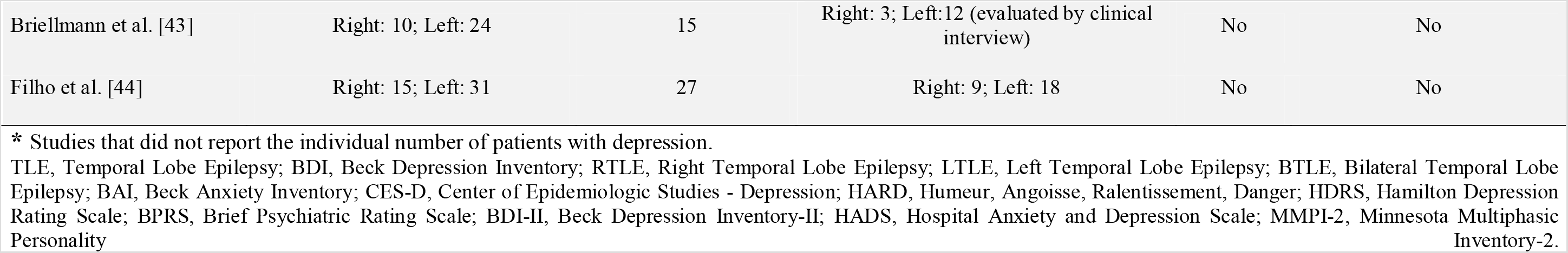
Clinical characteristics related to the lateralization of epileptic focus and depression and anxiety symptoms of non-surgical patients with Temporal Lobe Epilepsy.

## Discussion

To the date, studies have not yet completely clarified the links between the laterality of the epileptogenic area and behavioral symptoms and mood disorders. Although some authors reported a higher incidence of depression in left compared to right hemisphere damaged individuals [15, 28, 45–49, 39], some other authors also found no relationship between depressive symptoms and lateralization of the epileptic focus [50–52, 37]. The present systematic review showed no relation between the epileptic hemisphere and incidence of mood disorders.

The lack of standardized definitions of mood disorder may be a contributing factor to the negative outcome in an attempt to correlate mood disorder with the side of epilepsy involvement. To assess the parameters for defining cognitive functions and mood disorders, such as anxiety and depression, studies ranged from clinical interviews to different scales with conflicting cutoffs. This heterogeneity of data narrows the possibilities of inferring about the correlation of the lateralization of epileptic foci with the manifestation of psychological disturbances.

Mendez and colleagues suggested an association between depression and epileptogenic focus in the left hemisphere [14]. Nevertheless, Kalinin and Polyanskiy suggested that in patients with TLE the depression symptoms were determined by the right-sided focus while the anxiety symptoms were defined by the left-sided focus [53]. Also, an interesting study demonstrated that TLE patients with epileptic focus on the right hemisphere, presenting depression and anxiety as a solid syndrome, while TLE patients with focus on the left hemisphere, appear to have depression and anxiety as two independent syndromes [54]. However, Manchanda et al did not find correlation between epileptogenic focus laterality and depression development [55].

Some authors suggest that hippocampal dysfunction may be one of the leading factors in the development of depression, rather than the frequency of seizure or degree of disability [11–13, 41, 56]. In 2011, Gonçalves and Cendes demonstrated that patients with longer duration of epilepsy showed an increased risk of depression, however there was no association between seizure frequency and depression [32].

Briellmann et al. [43], however, found no correlation between hippocampal abnormalities with the prevalence of depression when using DSM-IV for diagnosis. Among their findings, changes in the amygdala were identified as a predictor of mood disorders. Similarly, three other studies point to amygdala volume as an associated parameter for mood disorders in epileptic patients [24, 27, 57]. Moreover, hippocampal atrophy is a common characteristic of TLE. Nevertheless, some studies showed no association between depression and the atrophy of epileptic hippocampus [43, 57].

Some other neurobiological factors should also be taken into consideration since epileptogenic focus laterality could not explain by itself the probability of depressive symptoms. Studies elucidated that other factors, such as motor lateralization and premorbid personality traits are considered important contributors for psychiatric disorders development in patients with TLE [58]. Moreover, cognitive impairment occurs frequently in patients with epilepsy and depression can influence the worsening of it. Paradiso and colleagues demonstrated that patients with TLE and comorbid depression performed significantly poorer results in several cognitive measures [59]. In the same context, Galioto and coauthors evidenced that depressive symptoms may contribute to executive impairments in patients with TLE [60]. Also, the antiepileptic drugs should be taken into consideration for associating mood disorders development in epileptic patients. Since, some types of AEDs are associated to beneficial effects for mood disorders while others are related with occurrence of depression symptoms [61]. The major limitation of this study is the risk of information bias because the researchers used different non-comparable methods for defining both cognitive functions and the presence or absence of mood disorders. In addition, older studies were included in order to cover all publications on the subject, but these were published previously to the creation of quantitative scales for mood evaluation. Furthermore, authors did not differentiate the degree of mood dysfunction and lateralization of epilepsy on a per patient basis. Detailed descriptions of patients’ therapeutic regimens were hardly found, another possible confounding factor at the time of data analysis. We encourage researchers to focus their efforts on well-established assessments such as the use of scales to define mood disorders and cognitive functions as well as, if possible, the joint analysis of seizure image, electroencephalogram and clinical pattern data, in order to establish the focus of epilepsy. Obviously, some other factors in addition to epileptogenic focus laterality should be considered important as contributors to the development of mood disorders in patients with epilepsy. In addition, the adequate description of the therapeutic scheme as well as socioeconomic and educational characteristics are important in the assessment of patients with mood disorders.

## Conclusions

This study shows mood disorders are prevalent in epileptic patients undergoing clinical treatment. However, to date there is no clear correlation between lateralization of epilepsy and the prevalence of mood disorders or cognitive impairment. Besides, the duration of epilepsy, type of seizures, the impact of antiepileptic and antidepressant drugs, cognitive dysfunction and premorbid personality traits need to be taken into consideration to better evaluate the relationship between symptoms of depression and anxiety and epilepsy. Well-conducted studies are needed to establish the correlation between the epilepsy lateralization and mood disorders in patients with refractory TLE.

## Data Availability

Not applicable.

## Acknowledgements

JCC is funded by CNPq (research productivity scholarship). JCC is supported by Conselho Nacional de Desenvolvimento Científico e Tecnológico – (CNPq) Brazil grant PQ 307372/2015–4. Our study was supported by the following grants: CNPq (Conselho Nacional de Desenvolvimento Científico e Tecnológico); Coordenação de Aperfeiçoamento de Pessoal de Nível Superior (CAPES).

## Abreviations

TLE: Temporal Lobe Epilepsy
BDI: Beck Depression Inventory
STAI: State Trait Anxiety Inventory
RTLE: Right Temporal Lobe Epilepsy
LTLE: Left Temporal Lobe Epilepsy
BTLE: Bilateral Temporal Lobe Epilepsy
AEDS: Antiepileptic Drugs
BAI: Beck Anxiety Inventory
CES-D: Center of Epidemiologic Studies – Depression
WCST: Wisconsin Card Sorting Test
WAIS-R: Wechsler Adult Intelligence Scale-Revised
LTL: Left Temporal Lobe
HARD: Humeur, Angoisse, Ralentissement, Danger
DSM-III-R: Diagnostic and Statistical Manual of Mental Disorders-III-Revised
HDRS: Hamilton Depression Rating Scale
NDI: Newcastle Depression Inventory
LPD: Levine-Pilowsky Depression Questionnaire
SCID: Structured Clinical Interview for DSM
BPRS: Brief Psychiatric Rating Scale
BDI-II: Beck Depression Inventory-II
QOLIE: Quality of Life in Epilepsy
CVLT-II: California Verbal Learning Test-II
WMS-III: Wechsler Memory Scale-III
D-KEFS: Delis–Kaplan Executive Function System
CWI: Colour-Word Interference Test
TMT-B: Trail-Making Test-B
DSM-IV: Diagnostic and Statistical Manual of Mental Disorders-IV
SDAS-21: Social Dysfunction and Aggression Scale
SCL-90-R: Symptom Checklist 90 Revised
WAIS-III: Wechsler Adult Intelligence Scale-Revised-III
MWT-B: Mehrfachwahl Wortschatz Test
HADS: Hospital Anxiety and Depression Scale
NART-R: National Adult Reading Test-Revised
WMS-R: Wechsler Memory Scale-Revised
MMPI-2: Minnesota Multiphasic Personality Inventory-2
BNT: Boston Naming Test
DCS-R: Diagnostikum fur Cerebralscha digung
VLMT: Verbal Learning and Memory Test
QOL: Quality of Life
POMS: Profiles of Mood States
COWA: Controlled Oral Word Association Test
BIS-11: Barratt Impulsiveness Scale 11
HAM-A: Hamilton Rating Scale for Anxiety
ICD-10: International Classification of Diseases-10
HAM-D: Hamilton Rating Scale for Depression
TMT: Trail Making Test
HADS-A: Hospital Anxiety and Depression Scale-Anxiety Subscale
MRI: Magnetic resonance imaging

## Declarations

### Ethics approval

Not applicable.

### Consent to participate

Not applicable.

### Consent for publication

Not applicable.

### Availability of data and material

Not applicable.

### Code availability

Not applicable.

## References

1. Téllez-Zenteno JF, Hernández-Ronquillo L (2012) A review of the epidemiology of temporal lobe epilepsy. Epilepsy Res Treat 630853. doi:10.1155/2012/630853

2. Pascual MRQ (2007) Temporal lobe epilepsy: clinical semiology and neurophysiological studies. Semin Ultrasound CT MR 28:416–23.

3. Spencer SS (2002) When should temporal-lobe epilepsy be treated surgically? The Lancet Neurology 1: 375–82.

4. Kanner AM (2006) Depression and epilepsy: a new perspective on two closely related disorders. Epilepsy Curr 6:141–146.

5. Bragatti JA, Torres CM, Londero RG, Martin KC, Souza ACD, Hidalgo MPL, et al. (2011) Prevalence of psychiatric comorbidities in temporal lobe epilepsy in a Southern Brazilian population. Arquivos de neuro-psiquiatria 69:159–65.

6. Dalmagro CL, Velasco TR, Bianchin MM, Martins APP, Guarnieri R, Cescato MP, et al. (2012) Psychiatric comorbidity in refractory focal epilepsy: a study of 490 patients. Epilepsy & Behavior 25: 593–97.

7. Edeh J and Toone B (1987) Relationship between interictal psychopathology and the type of epilepsy: results of a survey in general practice. The British Journal of Psychiatry 151: 95–101.

8. Schraegle WA and Titus JB (2017) The relationship of seizure focus with depression, anxiety, and health-related quality of life in children and adolescents with epilepsy. Epilepsy & Behavior 68: 115–22.

9. Shamim S, Hasler G, Liew C, Sato S, Theodore WH (2009) Temporal lobe epilepsy, depression, and hippocampal volume. Epilepsia 50: 1067–1071.

10. Lothe A, Didelot A, Hammers A, Costes N, Saoud M, Gilliam F, Ryvlin P (2008) Comorbidity between temporal lobe epilepsy and depression: a [18 F. MPPF PET study. Brain 131: 2765–82.

11. Gilliam FG, Maton BM, Martin RC, Sawrie SM, Faught RE, Hugg JW et al. (2007) Hippocampal 1HMRSI correlates with severity of depression symptoms in temporal lobe epilepsy. Neurology 68: 364–68.

12. Kondziella D, Alvestad S, Vaaler A, Sonnewald U (2007) Which clinical and experimental data link temporal lobe epilepsy with depression?. J Neurochem. 103: 2136–2152. doi:10.1111/j.1471-4159.2007.04926.x

13. Zanirati G, Azevedo PN, Venturin GT et al. (2018) Depression comorbidity in epileptic rats is related to brain glucose hypometabolism and hypersynchronicity in the metabolic network architecture. Epilepsia. 00:1–12. https://doi.org/10.1111/epi.14057

14. Mendez MF, Taylor JL, Doss RC, Salguero P (1994) Depression in secondary epilepsy: relation to lesion laterality. Journal of Neurology, Neurosurgery & Psychiatry 57: 232–33.

15. Altshuler LL, Devinsky O, Post RM, Theodore W (1990) Depression, anxiety, and temporal lobe epilepsy: laterality of focus and symptoms. Archives of Neurology 47: 284–288.

16. Kwon OY and Park SP (2014) Depression and anxiety in people with epilepsy. Journal of clinical neurology 10: 175–188.

17. Higgins J and Thomas J (2011) Cochrane handbook for systematic reviews of interventions.

18. Moher D, Liberati A, Tetzlaff J, Altman DG; PRISMA Group (2009) Preferred reporting items for systematic reviews and meta-analyses: the PRISMA statement. PLoS Med 6: e1000097.

19. Ehrlich T, Reyes A, Paul BM, Uttarwar V, et al. (2018) Beyond depression: The impact of executive functioning on quality of life in patients with temporal lobe epilepsy. Epilepsy Res 149: 30–36.

20. Reilly RE, Bowden SC, Bardenhagen FJ, Cook MJ (2006) Equality of the psychological model underlying depressive symptoms in patients with temporal lobe epilepsy versus heterogeneous neurological disorders. J Clin Exp Neuropsychol 28:1257–71.

21. Kohler C, Norstrand JA, Baltuch G, O’Connor MJ, Gur RE, French JA, Sperling MR (1999) Depression in temporal lobe epilepsy before epilepsy surgery. Epilepsia 40: 336–340.

22. Orjuela-Rojas JM, Martínez-Juárez IE, Ruiz-Chow A, Crail-Melendez D (2015) Treatment of depression in patients with temporal lobe epilepsy: A pilot study of cognitive behavioral therapy vs. selective serotonin reuptake inhibitors. Epilepsy Behav 51: 176–81. doi: 10.1016/j.yebeh.2015.07.033.

23. Robertson MM, Channon S, Baker J (1994) Depressive symptomatology in a general hospital sample of outpatients with temporal lobe epilepsy: a controlled study. Epilepsia 35: 771–7.

24. Tebartz Van Elst L, Woermann FG, Lemieux L, Trimble MR (1999) Amygdala enlargement in dysthymia – A volumetric study of patients with temporal lobe epilepsy. Biol Psychiatry 46: 1614–23.

25. Hermann BP, Seidenberg M, Bell B, Woodard A, Rutecki P, Sheth R (2000) Comorbid psychiatric symptoms in temporal lobe epilepsy: association with chronicity of epilepsy and impact on quality of life. Epilepsy Behav 1: 184–90.

26. Quiske A, Helmstaedter C, Lux S, Elger CE (2000) Depression in patients with temporal lobe epilepsy is related to mesial temporal sclerosis. Epilep Res 39:121–125.

27. Tebartz van Elst L, Woermann F, Lemieux L, Trimble MR (2000) Increased amygdala volumes in female and depressed humans. A quantitative magnetic resonance imaging study. Neurosci Lett 281: 103–6.

28. Piazzini A, Canevini MP, Maggiori G, Canger R (2001) Depression and anxiety in patients with epilepsy. Epilepsy Behav 2: 481–8.

29. Pulsipher DT, Seidenberg M, Jones J, Hermann B (2006) Quality of life and comorbid medical and psychiatric conditions in temporal lobe epilepsy. Epilepsy Behav 9: 510–4.

30. Tracy JI, Lippincott C, Mahmood T, Waldron B, Kanauss K, Glosser D, Sperling MR (2007) Are depression and cognitive performance related in temporal lobe epilepsy? Epilepsia 48: 2327–35.

31. de Oliveira GN, Kummer A, Salgado JV, Filho GM, David AS, Teixeira AL (2011) Suicidality in temporal lobe epilepsy: measuring the weight of impulsivity and depression. Epilepsy Behav 22: 745–9. doi: 10.1016/j.yebeh.2011.09.004.

32. Gonçalves EB, Cendes F (2011) Depression in patients with refractory temporal lobe epilepsy. Arq Neuropsiquiatr 69: 775–7.

33. Sanchez-Gistau V, Sugranyes G, Baillés E, et al (2012) Is major depressive disorder specifically associated with mesial temporal sclerosis? Epilepsia 53: 386–392.

34. Galioto R, Tremont G, Blum AS, LaFrance WC Jr, Crook CL, Davis JD (2017) Depressive Symptoms Contribute to Executive Deficits in Temporal Lobe Epilepsy. J Neuropsychiatry Clin Neurosci 29: 135–141.

35. Gonçalves EB, de Oliveira Cardoso TAM, Yasuda CL, Cendes F (2018) Depressive disorders in patients with pharmaco-resistant mesial temporal lobe epilepsy. J Int Med Res 46: 752–760.

36. Zingano BL, Guarnieri R, Diaz AP, Schwarzbold ML, Wolf P, Lin K, Walz R (2019) Hospital Anxiety and Depression Scale-Anxiety subscale (HADS-A) and The State-Trait Anxiety Inventory (STAI) accuracy for anxiety disorders detection in drug-resistant mesial temporal lobe epilepsy patients. J Affect Disord 246: 452–457.

37. Hermann BP, Seidenberg M, Haltiner A, Wyler AR (1991) Mood state in unilateral temporal lobe epilepsy. Biol Psychiatry 30: 1205–18.

38. Septien L, Gras P, Giroud M, Didi-Roy R, Brunotte F, Pelletier JL, Dumas R (1993) Depression and temporal epilepsy. The possible role of laterality of the epileptic foci and of gender. Neurophysiol Clin 23: 327–36.

39. Seidenberg M, Hermann BP, Noe A, Wyler AR (1995) Depression in temporal lobe epilepsy: interaction between laterality of lesion and Wisconsin Card Sorting Performance. Neuropsychiatry Neuropsychol Behav Neurol 8: 81–7.

40. Moore PM and Baker GA (2002) The neuropsychological and emotional consequences of living with intractable temporal lobe epilepsy: implications for clinical management. Seizure 11: 224–30.

41. Dulay MF, Schefft BK, Fargo JD, Privitera MD, Yeh H (2004) Severity of depressive symptoms, hippocampal sclerosis, auditory memory, and side of seizure focus in temporal lobe epilepsy. Epilepsy Behav 5: 522–31.

42. Helmstaedter C, Sonntag-Dillender M, Hoppe C, Elger CE (2004) Depressed mood and memory impairment in temporal lobe epilepsy as a function of focus lateralization and localization. Epilepsy Behav 5: 696–701.

43. Briellmann RS, Hopwood MJ, Jackson GD (2007) Major depression in temporal lobe epilepsy with hippocampal sclerosis: clinical and imaging correlates. J Neurol Neurosurg 2007; 78: 1226–30.

44. de Araújo Filho GM, Martins DP, Lopes AM, de Jesus Brait B, Furlan AER, Oliveira CIF, Marques LHN, Souza DRS, de Almeida EA (2018) Oxidative stress in patients with refractory temporal lobe epilepsy and mesial temporal sclerosis: Possible association with major depressive disorder? Epilepsy Behav 80: 191–196.

45. Robinson RG, Downhill JE (1995) Lateralisation of psychopathology in response to focal brain injury. In: Brain Asymmetry (Eds R. J. Davidson and K. Hugdahl). Cambridge, MA, USA, Mit Press 735.

46. Perini G, Mendius R (1984) Depression and anxiety in complex partial seizures. J Nerv Ment Dis 1984; 172: 287–90.

47. Strauss E, Wada J, Moll A (1992) Depression in male and female subjects with complex partial seizures. Arch Neurol 349: 391–2.

48. Carrieri PB, Provitera V, Iacovitti B, Iachetta C (1993) Mood disorders in epilepsy. Acta Neurol 15: 62–7.

49. Ring HA, Moriarty J, Trimble MR (1998) A prospective study of the early postsurgical psychiatric associations of epilepsy surgery. J Neurol Neurosurg Psychiatry 64: 601–4.

50. Naugle RI, Rodgers DA, Stagno SJ, Lalli J (1991) Unilateral temporal lobe epilepsy: an examination of psychopathology and psychosocial behavior. J Epilepsy 4: 164–7.

51. Lehrner J, Kalchmayr R, Serles W, et al. (1999) Health related quality of life, activity of daily living and depressive mood disorder in temporal lobe epilepsy patients. Seizure 8: 88–92.

52. Matthews WS and Klove H (1968) MMPI performances in major motor, psychomotor and mixed seizure classifications of known and unknown etiology. Epilepsia 9: 43–53.

53. Kalinin VV and Polyanskiy DA (2009) Focus laterality and interictal psychiatric disorder in temporal lobe epilepsy. Seizure 18: 176–9.

54. Kalinin VV, Zheleznova EV, Sokolova LV, Zemlyanaya AA, Subbotin K (2016) Focus Laterality and Interface Between Depression and Anxiety in Patients with Temporal Lobe Epilepsy.

55. Manchanda R, Schaefer B, McLaclan S, Blume WT, Wiebe S, Girvin JP, Parrent A, Derry PA (1996) Psychiatric disorders in candidates for surgery for epilepsy. J Neurol Neurosurg Psychiatry 61:82–89.

56. Peng W, Mao L, Yin D, Sun W, Wang H, Zhang Q, et al (2018) Functional network changes in the hippocampus contribute to depressive symptoms in epilepsy. Seizure 60: 16–22.

57. Richardson EJ, Griffith HR, Martin RC, Paige AL, Stewart CC, Jones J, Hermann BP, Seidenberg M (2007) Structural and functional neuroimaging correlates of depression in temporal lobe epilepsy. Epilepsy and Behavior. 10: 242–249.

58. Kalinin VV, Zemlyanaya AA, Zheleznova EV and Sokolova LV (2014) Neurobiological and Personality Risk Factors for Development of Obsessive-Compulsive Disorder in Patients with Epilepsy. Obsessive-Compulsive Disorder – The Old and the New Problems.

59. Paradiso S, Hermann BP, Blumer D, Davies K, Robinson RG (2001) Impact of depressed mood on neuropsychological status in temporal lobe epilepsy. J Neurol Neurosurg Psychiatry 70:180–185. doi:10.1136/jnnp.70.2.180

60. Galioto R, Tremont G, Blum AS, LaFrance WC Jr, Crook CL, Davis JD (2017) Depressive Symptoms Contribute to Executive Deficits in Temporal Lobe Epilepsy. J Neuropsychiatry Clin Neurosci. 29: 135–141.

61. Mula M and Schmitz B (2009) Depression in Epilepsy: Mechanisms and Therapeutic Approach. Ther Adv Neurol Disord 2: 337–344. doi: 10.1177/175628560933734

